# Cognitive profile partly mediates allergy-associated decrease in mental health

**DOI:** 10.1101/2020.02.01.20019778

**Authors:** Edwin S. Dalmaijer, Camilla L. Nord, Giacomo Bignardi, Alexander L. Anwyl-Irvine, Roma Siugzdaite, Tess A. Smith, Stepheni Uh, Addison Billing, Duncan E. Astle

**Affiliations:** MRC Cognition and Brain Sciences Unit, University of Cambridge, UK, 15 Chaucer Road, Cambridge, CB2 7EF, United Kingdom; Institute for Cognitive Neuroscience, University College London, UK, 17-19 Queen Square, London, WC1N 3AZ, United Kingdom

## Abstract

**Background:** There is a well-established link between allergies and mental health. While both atopic and food allergies are associated with increased levels of depression and anxiety, it is unclear what the mechanism behind this relationship is. Several theories touch upon potential psychological mechanisms, but until now putative cognitive mechanisms of the link between mental health and allergies have been unexplored.

**Methods:** We employed tablet-based deep cognitive phenotyping, and also recorded mental health, socio-economic status, and allergies in 533 children aged 7-9 years. We then employed mediation analyses to test whether cognitive mechanisms mediated the association between allergies and mental health.

**Findings:** In line with previous research, we found children with allergies reported higher levels of anxiety and depression. Furthermore, compared to children without allergies, they showed faster processing speed, equal verbal short-term memory, and worse performance on tests of fluid reasoning, number sense, search organisation, and spatial short-term memory. We confirmed that these variables predicted allergic state using logistic regression. Most importantly, we found that cognition partially mediated the relationship between allergy and both anxiety and depression.

**Interpretation:** Our results suggest that allergies bias children towards particular cognitive profiles, which in turn are risk factors for anxiety and depression. This supports the view that early cognitive interventions could reduce the number of allergic children that develops comorbid psychiatric conditions.

**Funding:** This study was supported by grant TWCF0159 from the Templeton World Charity Foundation to DEA, and by the UK Medical Research Council.

## Research in context

### Evidence before this study

It has been known for several decades that children with atopic allergies have an increased likelihood of developing symptoms of depression and anxiety, and more recent work extends this into food allergies. While it was initially thought that poor mental health was a consequence of allergy-induced inflammation, it has since been found that depression can precede allergy. In addition, parallel lines of work have found general decreases in cognitive function as a consequence or cause of atopic allergy.

### Added value of this study

We discovered cognitive mechanisms that mediate the relationship between allergies and mental health. We show that children with allergy are more likely to perform worse on various cognitive tasks (e.g. fluid reasoning and spatial short-term memory), but score better on tests of processing speed. These cognitive mechanisms predict allergic state in multivariable logistic regression. Crucially, cognitive profile partly mediates the relationship between allergy and worse mental health; a relationship that is no longer statistically significant after accounting for cognition.

### Implications of all the available evidence

Our study suggest that allergies bias children towards a cognitive profile of faster but shallower information processing, which is a known risk factor for anxiety and depression. This provides support for the idea that early cognitive intervention in children with allergies could prevent the development of mental health problems.

## Introduction

There is a well-established link between atopic allergies (e.g. asthma, eczema, rhinitis) and depression (1,2) and anxiety (3). Food allergies have similarly been associated with increased anxiety (4,5), and potentially increased depressive symptoms (6).

The causal nature of this association is unknown. While some accounts attribute depressive symptoms to allergy-induced increases in inflammation and immunoglobulin E (IgE) (7), others show that depression can induce allergy (8). This aligns with bidirectional theories of mental well-being and inflammation (9). Similarly, anxiety in people with allergies could arise from maladaptive generalisation of adaptive vigilance for food-related information (10).

In an altogether separate line of work, allergies are also associated with poorer cognitive performance in children. Underperformance on cognitive tasks at age 4 predicts atopy at age 6 (11), and children with allergies have poorer cognitive and school outcomes (12). One possible mechanism for the link between allergy and mental health is changes in cognition. Alterations in “cold” (non-emotional) cognition are thought to play a key role in the aetiology of depression in those with genetic or early-life vulnerabilities (13). Thus, an allergy might predispose children to mental health difficulties via changes in cognitive processing. We hypothesised that allergy would be associated with specific cognitive changes in children, and that these would mediate any association between allergy and mental health.

To test this hypothesis, we employed a broad and precise assessment of cognition in children aged 7-9 years, and recorded both their allergies and sub-clinical symptoms of anxiety and depression.

## Methods

We developed a computerised assessment battery that contained well-established tests of number sense, executive function (search organisation) and processing speed, fluid reasoning, inhibition, and spatial and verbal short-term memory. It also contained age-appropriate (spoken) questionnaires for allergies, anxiety, depression, family affluence, and school liking. Participants’ postcodes were obtained from schools or parents to compute their income deprivation affecting children index (IDACI). We have previously demonstrated that our assessments are of good reliability and validity (14), and have described the relationships between measures elsewhere (15).

We collected 468 usable datasets in local schools, and 69 from children who visited our facility (total N=533). They were 7-9 years old, and had a mean z-transformed IDACI of −0.27 (SD=0.92, min=−2.01, max=2.29; higher values indicate higher levels of deprivation). 409 cases were complete, and we used 9-nearest-neighbour imputation on 107 cases with 1 missing value, 15 with 2, 2 with 3, and excluded 4 cases with over 5 (10%) missing values.

Informed consent was obtained from parents and schools, in addition to assent from participating children. The study was approved by the Cambridge Psychology Research Ethics Committee (PRE.2017.102), and conducted in accordance with the Declaration of Helsinki.

In the sample who visited the facility, we could validate the allergy self-report accuracy. Parents but not their children reported an allergy in only 2 cases, and children but not their parents in only 5 cases. A small number of children (7 with parent data; 59 in total) mistook the “other unlisted allergy” for the “no allergy” button, and were not included. In total, 206 children (39% of the sample) indicated they had one or more allergies, which is in line with population estimates (3,16).

Simple group differences between children with and without allergies were computed using Welch’s *t*-test, which is suitable for groups of different sizes and variances (degrees of freedom are dependent on variance, and thus vary between comparisons). We also employed multivariable logistic regression to predict allergic state from socio-economic, mental health, and cognitive variables; and estimated 95% confidence intervals using 3000 bootstrapping iterations, in each of which we used 5-fold cross-validation. Finally, mediation analyses (17) tested the relationship between allergies and mental health, and potential mediation through cognitive profile.

### Role of the funding source

The study sponsors, Templeton World Charity Foundation and UK Medical Research Council, had no involvement in the study design, data collection, analysis, interpretation, writing, or the decision to submit this manuscript for publication.

## Results

After confirming that children with allergies reported the expected higher levels of anxiety [*t*(410)=2.62, *p=*0.009, *d*=0.24] and depression [*t*(404)=3.11, *p=*0.002, *d*=0.28], we tested whether they showed a different cognitive profile (Figure 1A). Indeed, children with allergies scored poorer on tests of fluid reasoning [*t*(367)=3.66, *p*<0.001, *d*=0.34], number sense [*t*(453)=3.78, *p*<0.001, *d*=0.33], search organisation [*t*(453)=2.71, *p=*0.007, *d*=0.24], and spatial short-term memory [*t*(435)=2.32, *p=*0.021, *d*=0.21], but they also showed higher average processing speed [*t*(526)=−2.56, *p=*0.011, *d*=−0.20]. No differences were observed in school liking [*t*(424)=1.95, *p=*0.052], inhibition [*t*(440)=0.88, *p=*0.381], and verbal short-term memory [*t*(461)=−0.11, *p=*0.911]. We controlled for socio-economic status (SES; deprivation and affluence) in these analyses, but results were highly similar when we did not.

**Figure 1.**
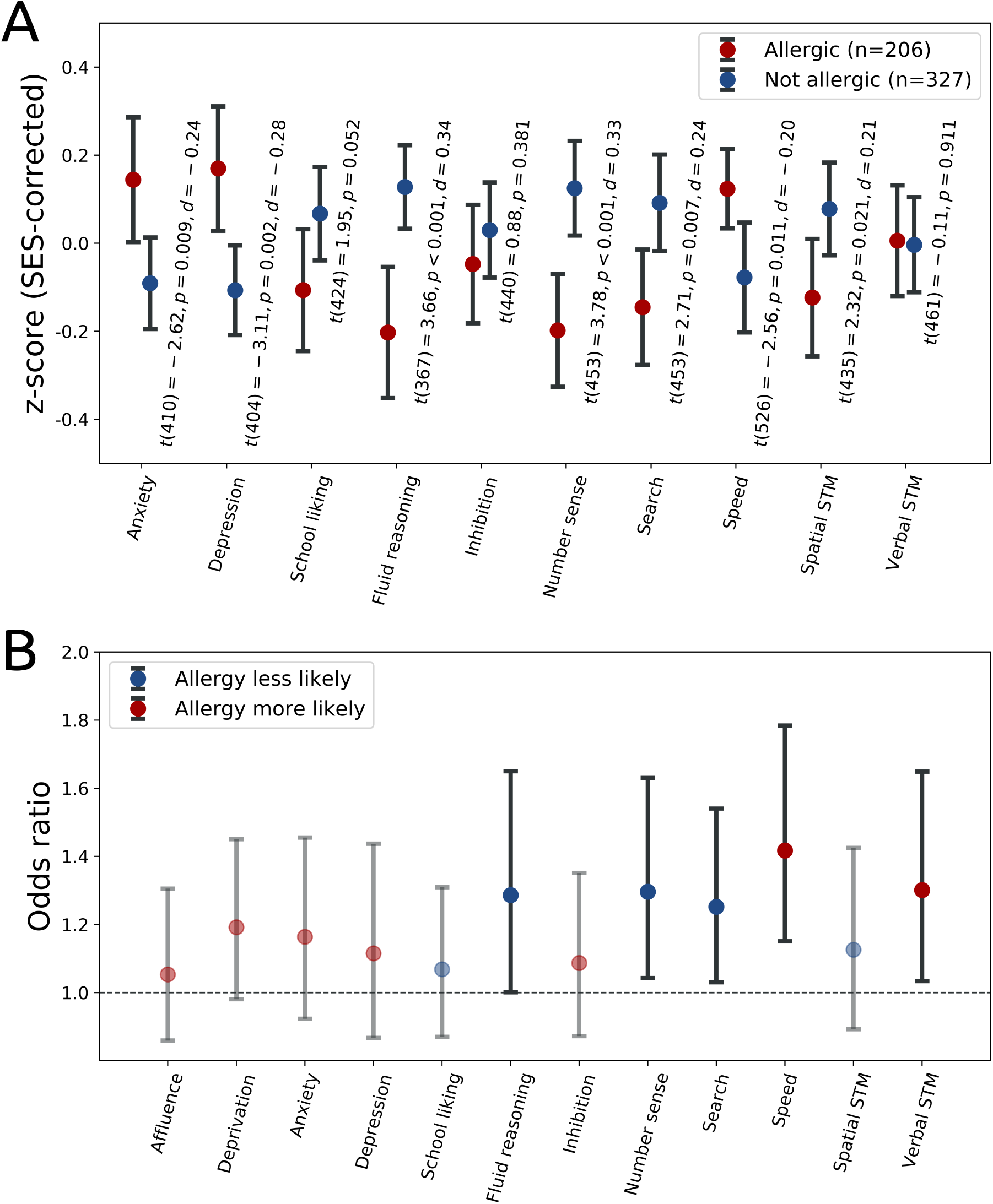
The top panel (A) shows differences between allergic and non-allergic children on cognitive tasks. Annotated scores are from Welch’s t-tests. The bottom panel (B) shows odds ratios and their 95% confidence intervals computed through 3000 bootstrapping iterations of logistic regression with 5-fold cross-validation. Blue markings indicate predictors of having no allergy, and red of having any allergy. Opaque markings indicate statistically significant predictors (odds ratio confidence interval > 1).

Next, we tested what variables could accurately predict whether children had an allergy using logistic regression. We found that cognitive profile predicted allergic state, but other factors (including mental health) did not. Having no allergies was predicted by better fluid reasoning [β=−0.25, CI=[−0.5, −0.001]; OR=1.29, CI=[1.00, 1.65]], number sense [β=−0.29, CI=[−0.49, −0.04]; OR=1.30, CI=[1.04, 1.63]], and search organisation [β=−0.22, CI=[−0.43, −0.03]; OR=1.25, CI=[1.03, 1.54]], while having any allergy was predicted by better processing speed [β=0.35, CI=[0.14, 0.58]; OR=1.42, CI=[1.15, 1.78]] and better-than-expected verbal short-term memory [β=0.26, CI=[0.03, 0.50]; OR=1.30, CI=[1.03,1.65]] (Figure 1B). Variables that did not contribute to predicting allergic state were affluence [β=0.05, CI=[−0.15, 0.27]], deprivation [β=0.18, CI=[−0.02, 0.37]], anxiety [β=0.15, CI=[−0.08, 0.37]], depression [β=0.11, CI=[−0.14,0.36]], school liking [β=−0.07, CI=[−0.27, 0.14]], inhibition [β=0.08, CI=[−0.14, 0.30]], and spatial short-term memory [β=−0.12, CI=[−0.35, 0.11]].

Finally, we tested whether different cognitive profiles could account for the increased mental health symptom burden in children with allergies. While allergies significantly predicted higher anxiety [R=0.11, *p*=0.008] and depression [R=0.14, *p*=0.002], this was partially mediated by cognitive profile (Figure 2) for anxiety [remaining direct effect c’=0.177 (0.089), *p*=0.046; mediation ab=0.058 (0.013), *p*<0.001, percentage mediated=24.66%] and depression [remaining direct effect c’=0.155 (0.087), *p*=0.076; mediation ab=0.121 (0.013), *p*<0.001, percentage mediated=43.90%]. Thus, the statistically significant direct relationship between allergies and mental health was rendered unconvincing (anxiety, *p*=0.046) or non-significant (depression, *p*=0.076) after accounting for cognitive profile. We controlled for SES in these analyses too, but results were again highly similar when we did not.

**Figure 2.**
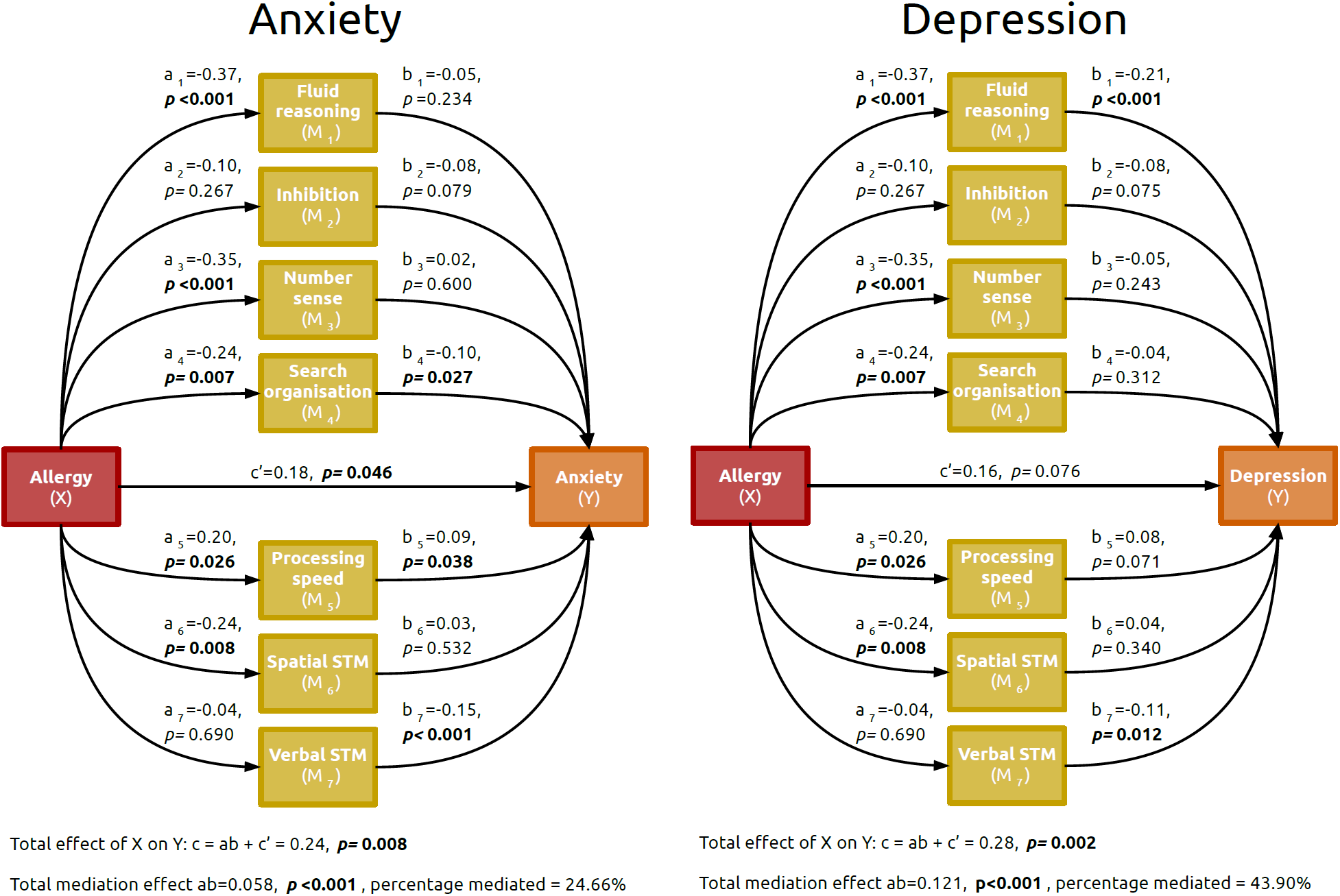
Mediation analyses for the relation between allergy (red box) and anxiety (orange box, left panel) and depression (orange box, right panel), with multiple cognitive factors as mediators (yellow boxes). Standardised coefficients and associated p values are reported for each relation, and the total and combined mediation effects are reported in the text below each model. The mental health measures reported here are corrected for socio-economic status (affluence and deprivation), but results are highly similar for uncontrolled outcomes (percentage mediated of 24.97 and 41.93, respectively).

## Discussion

We measured cognitive processing and mental health in a large cross-sectional sample of children. Unusually, we included a wide breadth of cognitive measures, representing a deep cognitive phenotyping assessment. This methodological strength allowed us to more fully characterise the profiles of children with and without allergies than previous studies in the literature. We used this approach to answer two questions: the effect of allergy on cognitive profiles in children, and whether differences in cognitive profile could account for allergy-associated worsening of mental health. We found that children with allergies had worse fluid reasoning, number sense, and search organisation compared to their peers. However, children with allergies also showed faster processing speed, and no difference in verbal short-term memory. Replicating previous work (1–6), we found that children with allergies showed increased depression and anxiety symptoms, even after controlling for SES. However, this relationship was partly mediated and thereby rendered statistically unreliable when cognitive profile was included in the model. We characterised a specific cognitive profile associated with allergies in children: quicker processing speed, but compromised accuracy on higher-order cognitive tasks. Crucially, this cognitive profile partially mediated the relationship between allergies and mental health.

We suggest that having allergies can bias children towards a cognitive profile of faster but shallower processing of information, which in turn is a risk factor for anxiety (18) and depression (13). Our findings provide a cognitive mechanism that can explain why some reports found allergy onset to precede anxiety (10) or depression (11) symptoms.

Our work adds to a large literature on how allergy might induce changes in cognition. A prominent theory posits that adaptive adaptive vigilance for allergy-specific for allergy-related stimuli could bias attention to and interpretation of potentially threatening material (e.g. peanuts or pollen), and thereafter generalise to maladaptive global anxiety (10). However, the mechanism through which this maladaptive generalisation to global anxiety could originate remained unclear. One possibility is via changes in executive function and other “cold” cognitive measures, as we suggest here. In addition, allergy and certain cognitive profiles could share common biological mechanisms. For example, the G protein-coupled receptor 154 gene is associated with asthma and increased serum IgE, and also with HPA-axis responses to stress and anxiety (11), which could in turn shift both mental health and cognition (19). Further cognitive changes could arise from neurobiological consequences of being in an atopic state, for example its effects (through a shift from T helper 1 to 2 responses that produce interleukin-4) on the serotonin metabolism (2), which is crucial for cognitive flexibility (20,21).

It should be noted that, although no statistically significant direct relationship remains between allergy and depressive symptoms after accounting for cognitive variables, the mediation is only partial: our cognitive measures do not fully account for the association between allergy and mental health. The remaining variance could be explained by other cognitive or non-cognitive factors, and should be explored in the future. Furthermore, our results are based on cross-sectional data. Ideally, these variables would be measured in a large longitudinal sample to track the emergence of major mental health disorders that typically emerge in later ages than those included in our sample. Nevertheless, we report one major source of variance in allergy-associated worsening of mental health: an altered cognitive profile. Future work should explore why an altered cognitive profile represents such an important source of variance. Perhaps this cognitive profile instantiates a mental state more vulnerable to the effects of stress or other mental health risk factors.

Our findings highlight the potential for deep cognitive phenotyping to explain existing associations (here between allergy and mental health) described in the literature, and underscore the need for early intervention in children with allergies. As noted previously, early intervention in children with allergies can improve well-being in both parent and child, improve compliance and outcomes, and reduce mortality (22). Our results suggest that early intervention also has the potential to counteract effects on cognition, which may have further implications for mental health.

## Conclusion

Allergy-associated increases in depression and anxiety symptoms are mediated by a cognitive trade-off: Children with allergies are more likely to process information faster, while their peers without allergies process at greater depth. Accounting for cognitive differences substantially diminishes the relationship between allergic state and mental health, highlighting cognition’s central role in allergy-associated changes in mental health.

## Data Availability

The authors will make a synthetic dataset available with the publication of this manuscript, as well as the analysis code via GitHub (https://github.com/esdalmaijer/). It will be constructed to match the properties of the original data (using R package synthpop), without the danger of potential deanonymisation of participants. The original data will be available from the corresponding author upon request, and upon signing an agreement to not publicly share it. This ensures that our analyses are reproducible, while maintaining the privacy of our participants.

## Declaration of interests

The authors declare that there are no relevant financial or personal relationships that could be conceived as actual or potential conflicts of interests affecting the analysis, interpretation, or publication of the current study.

## Data sharing

The authors will make a synthetic dataset available with the publication of this manuscript, as well as the analysis code via GitHub (https://github.com/esdalmaijer/). It will be constructed to match the properties of the original data (using R package *synthpop*), without the danger of potential deanonymisation of participants. The original data will be available from the corresponding author upon request, and upon signing an agreement to not publicly share it. This ensures that our analyses are reproducible, while maintaining the privacy of our participants.

## Author contributions

Substantial contributions: ESD, GB, ALA-I, AB, and DEA conceptualised the design. ESD, GB, ALA-I, TAS, RS, and SU collected the data; ESD analysed the data; ESD, CLN, and DEA interpreted the data. Drafting and revising: ESD, CLN, and DEA drafted the manuscript; all authors provided critical feedback. Final approval and accountability: All authors approved of the final manuscript, and agreed to be accountable for the accuracy and integrity of the work.

## Notes

### Competing Interest Statement

The authors have declared no competing interest.

